# The role of viral interference in shaping RSV epidemics following the 2009 H1N1 influenza pandemic

**DOI:** 10.1101/2024.02.25.24303336

**Authors:** Ke Li, Deus Thindwa, Daniel M Weinberger, Virginia E Pitzer

## Abstract

**Background:** Disruptions in respiratory syncytial virus (RSV) activity were observed in different countries following the 2009 influenza pandemic. Given the limited use of non-pharmaceutical interventions, these disruptions do provide an opportunity to probe viral interference due to the out-of-season epidemics. The objectives of the study are twofold: to characterize atypical RSV activity in the United States (US) and to explore the mechanisms underlying changes in RSV epidemics following the pandemic.

**Methods:** Laboratory-confirmed RSV cases across 10 US regions from June 2007 to July 2019 were analyzed. A dynamic time warping method was used to characterize RSV activity in different seasons. A two-pathogen model was constructed to explore viral interference mechanisms. A sampling-importance resampling method was applied to estimate the effects of viral interference.

**Results:** We found that RSV activity was reduced following the influenza pandemic in the 2009/10 season across all regions in the US. By contrast, we found an enhanced but delayed RSV epidemic across the US in the 2010/11 season. Using a mathematical model, we identified three potential viral interference mechanisms that could explain the change of RSV activity following the pandemic. The pandemic influenza may interfere with RSV to reduce susceptibility to RSV coinfection, or shorten the RSV infectious period, or decrease RSV infectivity in co-infections.

**Conclusions:** This study provides statistical evidence for atypical RSV seasons following the influenza pandemic in the US and sheds light on viral interference mechanisms affecting RSV epidemics, offering a model-fitting framework for analyzing surveillance data at the population level.

## INTRODUCTION

Respiratory syncytial virus (RSV) infections are a major public health concern for infants and young children, causing severe lower respiratory tract infections [1]. In the United States (US) and other temperate regions, RSV activity is strongly seasonal, typically beginning in the fall and peaking in winter [2]. Out-of-season RSV epidemics were observed in the US following the COVID-19 pandemic, which was likely due to the implementation of non-pharmaceutical interventions (NPIs) to reduce the spread of SARS-CoV-2 [3,4]. However, disruptions to RSV activity were also observed following the 2009 influenza pandemic in different countries despite the limited implementation of NPIs [5–10]. Given the limited use of NPIs, the disruptions do provide an opportunity to probe viral interference due to the out-of-season epidemics.

While it is evident that viral interference occurs at the host level [11–13], demonstrating the presence of viral interference at the population level and quantifying its impact on disease transmission is more challenging. To date, most studies that analyzed population-level viral interference have primarily focused upon statistical associations between reported positive cases of different viruses, using regression and correlation analyses [13,14]. Neither the biological mechanisms underpinning potential viral interactions, nor the strength of interactions could be determined or quantified using these models.

Mathematical models that explicitly depict the underlying mechanisms of viral transmission have advantages in being able to integrate heterogeneous mechanisms and test different hypotheses [15,16]. In particular, mathematical models have been proposed to study the effect of viral interference between RSV and influenza viruses at the population level and to quantify the interactions by fitting the models to incidence data [17–19]. However, these models did not capture the natural infection history of RSV, which are characterized by intermediate immunity lying between perfectly and imperfectly immunizing infections. In this work, we started by analyzing laboratory-confirmed cases of RSV and the pdmH1N1 virus in different regions in the US. We applied a dynamic time warping and hierarchical clustering method to identify atypical RSV seasons following the pandemic. We then built a mechanistic, age-stratified mathematical model that incorporates an RSV transmission model into an influenza transmission model, coupled with hypothesized viral interference mechanisms. Using a sampling-importance resampling method to explore the parameter space, we simulated various scenarios for influenza dynamics and quantified the potential interactions between RSV and pdmH1N1 influenza virus at the population level.

## METHODS

### Laboratory reporting of RSV and influenza

Weekly data on laboratory reporting of RSV tests in 10 Health and Human Services (HHS) regions in the US from June 2007 to July 2019 were obtained from the National Respiratory and Enteric Virus Surveillance System. Positive RSV tests were detected using three diagnostic methods: 1) antigen detection; 2) reverse transcription polymerase chain reaction (RT-PCR); and 3) viral culture. Correspondingly, the data on laboratory reporting of influenza tests in the US from the same period were obtained from the Center for Disease Control and Prevention (CDC) website https://gis.cdc.gov/grasp/fluview/flu_by_age_virus.html.

The raw laboratory data were rescaled based on the number of positive tests to account for variations in testing practices over time [20]. We first calculated a one-year moving average of the weekly number of RSV or influenza tests (both positive and negative tests) in each region centered on each week. We then calculated a weekly scaling factor for each region equal to the average number of RSV or influenza tests during the entire period of reporting (i.e., 12 epidemic seasons from 2007-2019) divided by the one-year moving average. The rescaled number of RSV or influenza-positive tests for each region was then calculated as the reported number of positive tests multiplied by the weekly scaling factor.

### Demographic Data

Information about population size in each age group was obtained from the US Census Bureau’s American Community Survey. Birth rates varied between regions and over time based on the crude annual birth rate for each HSS region from 1990 to 2019. These were obtained from https://wonder.cdc.gov/controller/datarequest/D66. To capture aging among infants and children more accurately in our mathematical model, we divided the <1 year and 1-4 years age class into 12-month age groups. The remaining population was divided into 5 classes: 5-9 years, 10-19 years, 20-39 years, 39-60 years and >60 years old. Individuals were assumed to age exponentially into the next age class, with the rate of aging equal to the multiplicative inverse of the width of the age class. Data on age-specific contact rates were available from [21].

## RESULTS

### RSV activity before and following the 2009 influenza pandemic

RSV epidemics exhibit consistent seasonal timing and duration in the US, with variation in timing between regions, typically starting in the fall and peaking in the winter (**Fig. 1A** and **SFig. 1A**). The influenza pandemic (the shaded area, **Fig. 1A** and **SFig. 1A**) began in April 2009, after the 2008/09 RSV season. This was followed by a second wave in most regions that started at the end of 2009, before the peak of the 2009/10 RSV season.

**Fig. 1.**
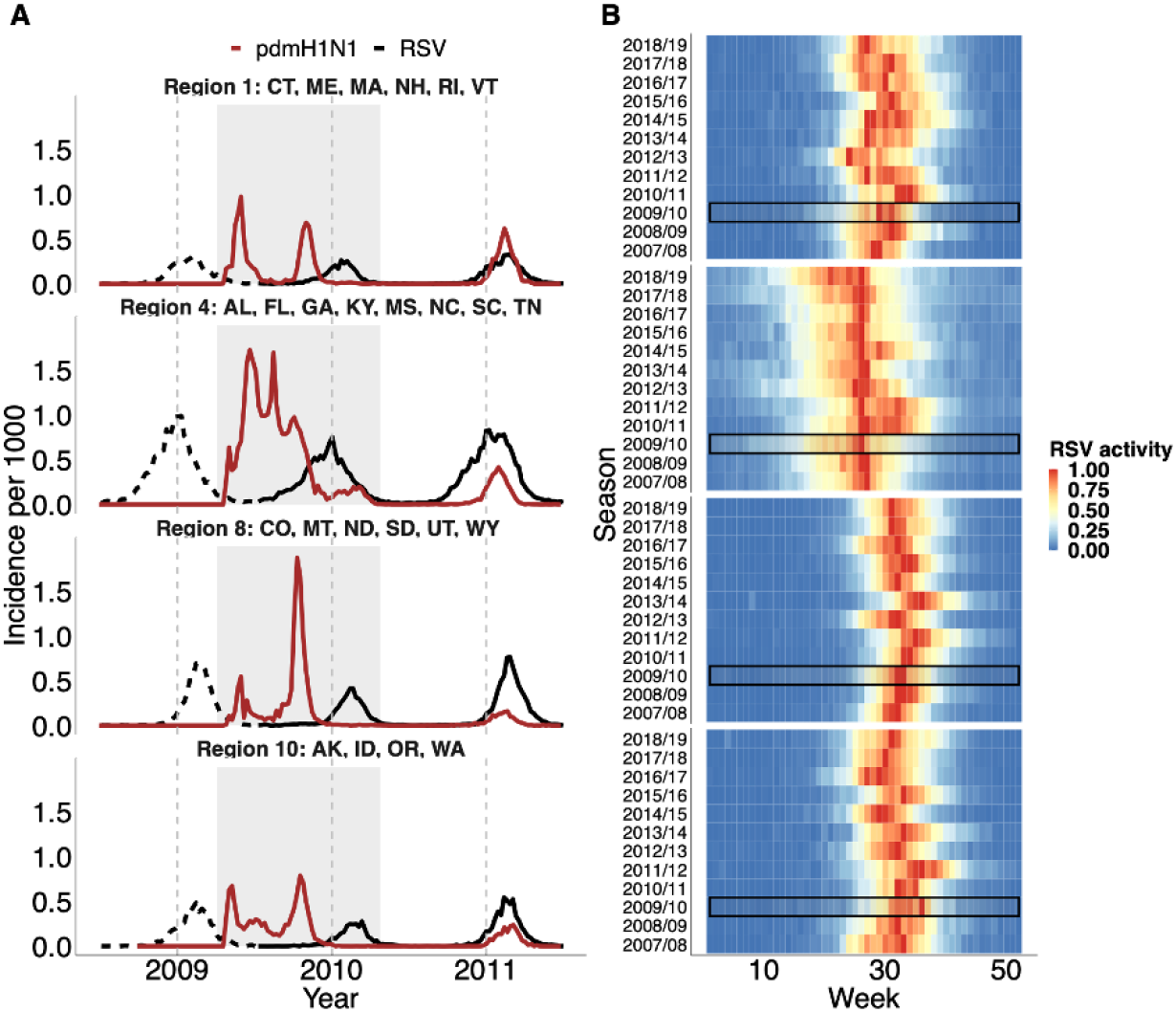
Laboratory-confirmed positive tests for pdmH1N1 virus and RSV following the pandemic. **(A)** Positive laboratory tests for pdmH1N1 virus (red) and RSV (black) before (dashed line), during (shaded area) and following the 2009 influenza pandemic in Region 1 (CT: Connecticut, ME: Maine, MA: Massachusetts, NH: New Hampshire, RI: Rhode Island, VT: Vermont), Region 4 (AL: Alabama, FL: Florida, GA: Georgia, KY: Kentucky, MS: Mississippi, NC: North Carolina, SC: South Carolina, TN: Tennessee) Region 8 (CO: Colorado, MT: Montana, ND: North Dakota, SD: South Dakota, UT: Utah, WY: Wyoming) and Region 10 (AK: Alaska, ID: Idaho, OR: Oregon, WA: Washington) of the US. Dashed gray lines indicate the 1st of January each year. **(B)** Heatmap of RSV activity by epidemic season for the four selected regions. Epidemic seasons are defined as starting in July and ending in June of the following year. RSV activity was calculated as the fraction of positive tests among the total number of tests for each season. The resulting fraction was then normalized to a range between 0 and 1, with red indicating high activity and blue indicating low activity. RSV activity during the pandemic 2009/10 season is highlighted in the black box.

Based on the laboratory reports of RSV-positive specimens, we showed that Northeast and Southeast regions (e.g., regions 1 and 4) exhibited consistent annual patterns of RSV activity, with a steady onset and peak timing across different seasons (**Fig. 1B** and **SFig. 1B**). However, we noted delayed RSV activity in 2010/11 compared with other seasons in the two regions, as indicated by a shift of intense RSV activity to later epidemic weeks. Upper Midwest and Northwest regions (e.g., regions 8 and 10) exhibited biennial patterns, with RSV tending to start and peak earlier with a larger epidemic in even-numbered seasons (e.g., 2010/11) compared to the odd-numbered seasons (e.g., 2011/12). In these regions, we also observed delayed RSV activity in the 2010/11 compared to other even-numbered years. The delayed RSV epidemics were also found in other regions (**SFig. 1B**)

### Atypical RSV seasons following the influenza pandemic

To detect the differences in RSV activity across seasons, we then applied dynamic time warping (DTW) and a hierarchical clustering method to compare RSV activity following the influenza pandemic with that of other RSV seasons (see **Supplementary Text**). The hierarchical clustering results, based on the optimal alignments of RSV activity (**SFig. 2**), demonstrate the similarities of RSV epidemics across different epidemic seasons. In regions 1 and 4 where RSV exhibit an annual pattern, we observed that the 2009/10 and 2010/11 seasons were closer in the dendrogram, grouped into the same cluster, compared with other seasons before or after the pandemic (as shown in the dashed box, **Figs. 2A** and **B**). We also showed that even within the same cluster, the 2009/10 season was distant from the 2010/11 season, suggesting a different pattern between the two seasons following the pandemic. Notably, we found that the hierarchical clustering method successfully captured the biennial pattern in regions 8 and 10 (**Figs. 2C** and **D**) and other regions in the US (**Fig. S3**), grouping even seasons and odd seasons into different clusters. In particular, we found that the 2009/10 season was grouped with even-year seasons in Region 8, indicating unusual RSV activity. Overall, the clustering results demonstrate an atypical RSV activity in the 2009/10 and 2010/11 seasons following the pandemic.

**Fig. 2.**
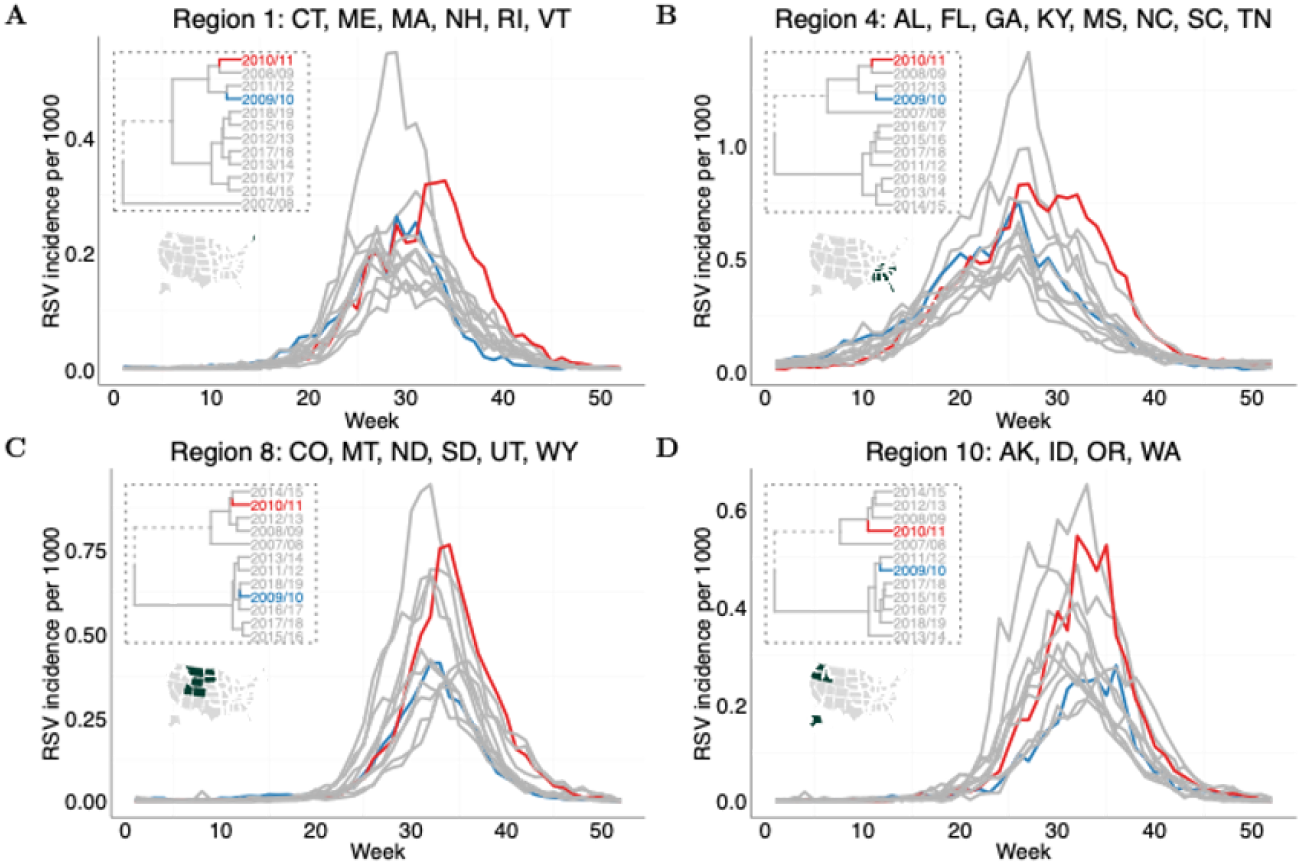
Hierarchical clusters and RSV time-series in the four selected regions. An agglomerative clustering algorithm with a Ward variance method was used to group the 12 RSV epidemic seasons in each region based on the alignment paths (**Fig. S2**). The dendrograms (in the dashed box) give the clustering results based on the optimal alignments between time-series computed by DTW. **(A)**-**(D)** show the dendrogram and corresponding RSV time-series in Region 1, 4, 8 and 10, respectively. RSV time-series following the pandemic seasons are highlighted in blue (2009/10) and red (2010/11). The clustering results for other regions are given in **Fig. S3**.

We then calculated the intensity and center of gravity of the RSV epidemic for each region and compared them in the seasons before and after the pandemic. We found that the peak timing of RSV activity in the 2009/10 season was similar with that of other seasons (**Fig. 3A**). However, all regions experienced decreased RSV activity in the 2009/10 season (**Fig. 3B**). Notice that all points are aligned above the diagonal line. By contrast, we observed delayed peaking time of RSV activity (**Fig. 3C**) with reduced RSV intensity in the 2010/11 season compared with other seasons (**Fig. 3D**).

**Fig. 3.**
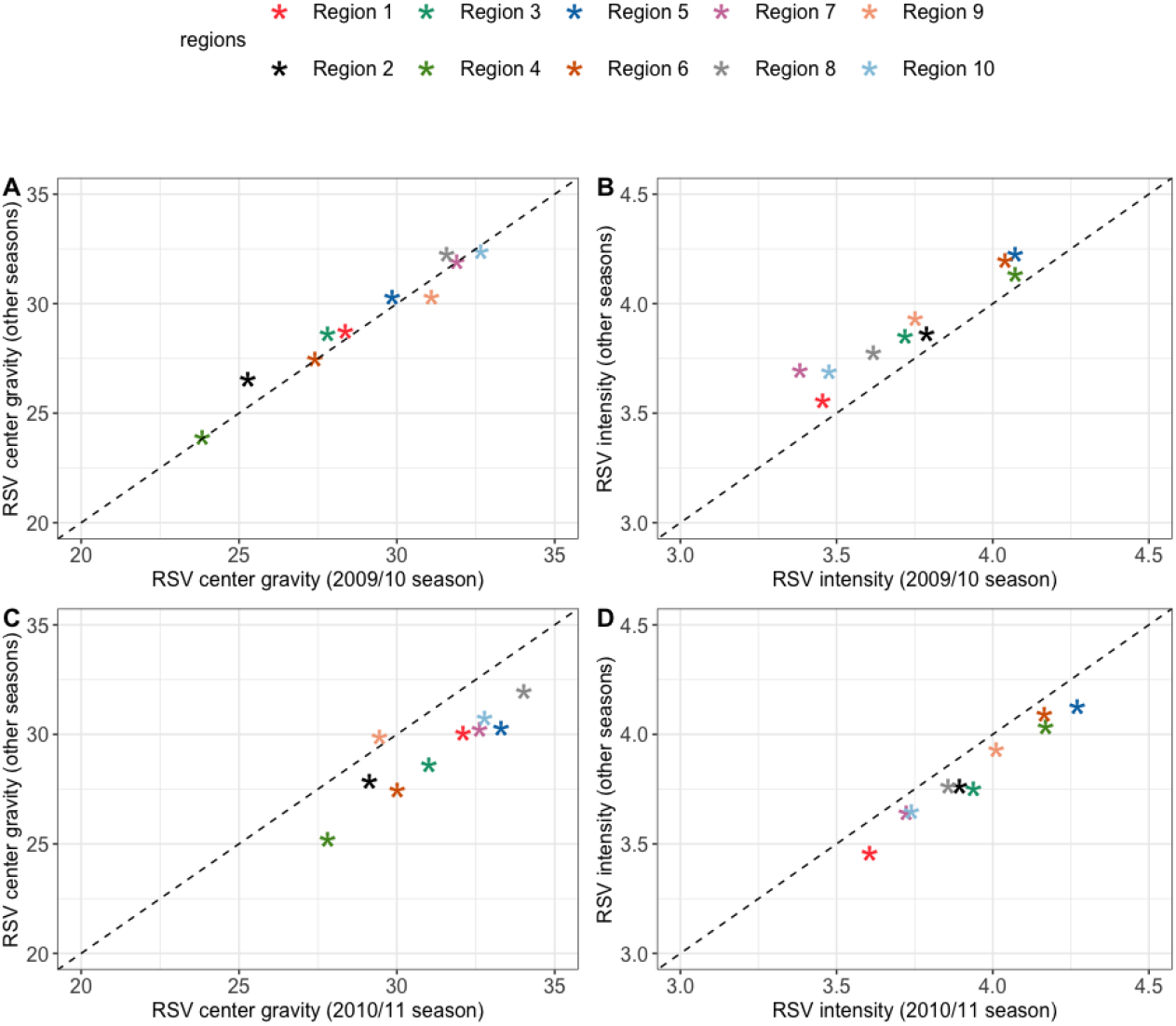
Timing and intensity of RSV activity. Comparison of the center of gravity of RSV activity in the (A) 2009/10 and (C) 2010/11 seasons with other seasons. The vertical axis indicates the median value of the center of gravity of RSV activity for seasons other than 2009/10 and 2010/11. For regions 7-10 where biennial RSV epidemics were observed, the vertical axis indicates the median value of all odd-numbered or all even-numbered seasons. Comparison of the intensity of RSV activity in the (B) 2009/10 and (D) 2010/11 seasons with other seasons.

### Transmission model analyses

Having demonstrated the changes of RSV activity following the influenza pandemic, we next examine whether the disrupted activity could be explained by the influenza pandemic using a mathematical model and statistical inference methods (see **Supplementary Text**). With the viral interference effects from influenza, the model was able to replicate the decreased RSV activity in the 2009/10 season and increased RSV activity in the 2010/11 season for all regions as observed (**Fig. 4**). We found strong correlations between the observed and predicted RSV intensity in the 2009/10 season (**Fig. 4A**, Pearson’s correlation coefficient *r* = 0.97, p < 0.001) and 2010/11 season (**Fig. 4B**, *r* = 0.95, p < 0.001) for all regions. Notably, we found that only the model, incorporating viral interference effects, could reproduce the biennial pattern observed following the pandemic in the Upper Midwest and Northwest regions (**Fig. 4C** and **SFig. 5**). However, our model could not reproduce the delayed RSV peak in the 2010/11 season, even when accounting for viral interference.

**Fig. 4.**
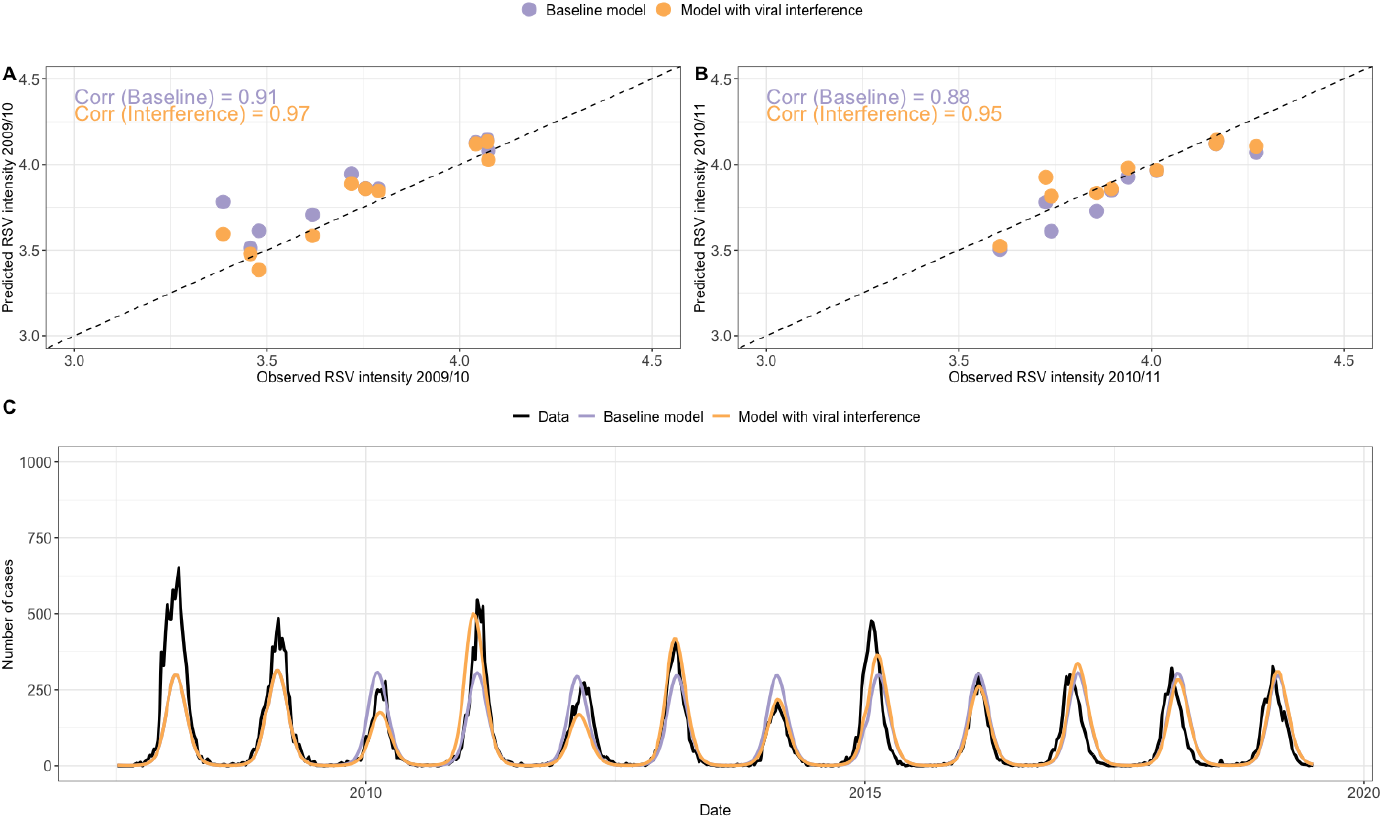
Model predictions of RSV activity. The correlation coefficients of RSV intensity between observed data and model predictions for the **(A)** 2009/10 and **(B)** 2010/11 season for all regions. Each point represents one region. **(C)** Model predictions of RSV epidemics in Region 10 where RSV activity exhibits a biennial pattern. In the baseline model, we assume no viral interference effects (i.e., θ = 1, ξ = 1 or η = 1). In the model with viral interference, we use median estimates of θ and set the other two effects to the baseline values. The model predictions with the other viral interference mechanisms are given in **SFig. 5**.

**Fig. 5.**
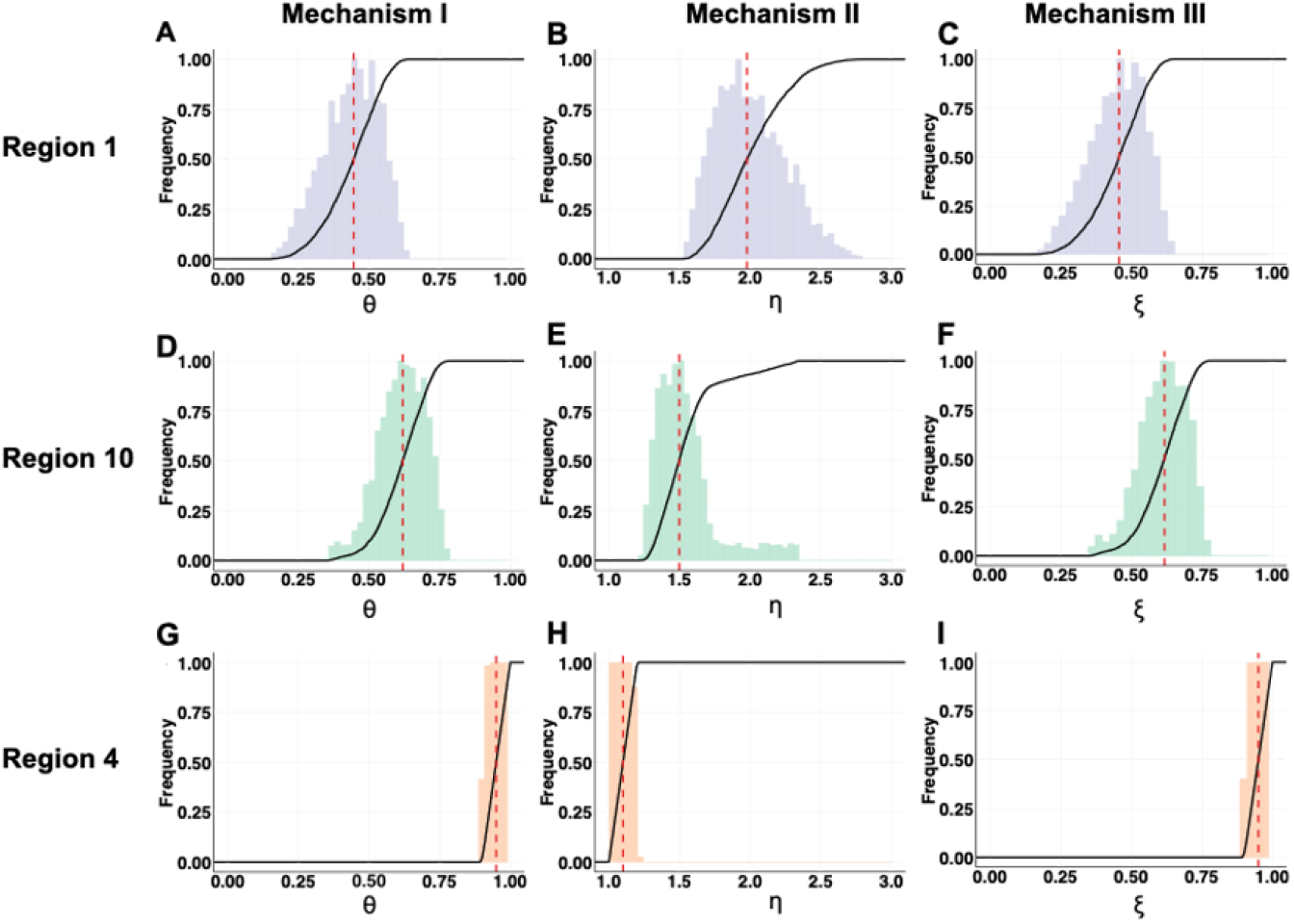
Identified parameter space for viral interference parameters. Histograms show the frequency of the values of viral interference parameters. The cumulative density functions (CDFs) are given by the solid lines, and the dashed red lines indicate the median estimates. **(A, D, G)** show the identified parameter distribution for the viral interference mechanism that reduces the host’s susceptibility to RSV infection in Region 1, 10 and 4, respectively. **(B, E, H)** show the identified parameter distribution for the viral interference mechanism that shortens RSV infectious period in Region 1, 10 and 4, respectively. **(C, F, I)** show the identified parameter distribution for the viral interference mechanism that reduces RSV infectivity in Region 1, 10 and 4, respectively.

We further extracted the marginal distributions for estimated viral interference parameters for in regions 1, 10, and 4, where RSV activity exhibits annual, biennial patterns and the earliest onset in the US, respectively. We found that the median estimate for the reduction of host susceptibility to RSV infection when infected with influenza virus (θ) was 0.44 (95% credible interval (CI): 0.23-0.60, **Fig. 5A**). In epidemiological terms, the median estimate for θ indicates that the presence of the pdmH1N1 infection reduces the likelihood of hosts being subsequently infected with RSV by nearly 60%. The median estimate for η implies the presence of the pdmH1N1 co-infection halves the RSV infectious period, i.e. increases the rate of recovery by a factor 1.98 (95% CI: 1.62-2.53, **Fig. 5B**). Similarly, the median estimate for ξ was 0.46 (95% CI: 0.20-0.80, **Fig. 5C**), suggesting the pdmH1N1 infection reduces RSV infectivity by 53%. We found comparable viral interference effects in Region 10 (**Figs. 5D-F**). By contrast, we found weak interference effects between the viruses in Region 4, such that all estimated parameters were close to 1 (**Figs. 5G-I**).

## DISCUSSION

By focusing on the dynamics of RSV following the 2009 H1N1 influenza pandemic, we found evidence supporting the presence of interactions between the viruses at the population level and examined the underlying mechanisms. Our results support and contribute to the current knowledge from several observational studies that RSV activity was disrupted following the pandemic [5–10]. Using a two-pathogen transmission model, we assessed potential interactions between the viruses and identified three mechanisms of viral interference that could replicate the relative difference of RSV activity in the two epidemic seasons following the pandemic. The identified parameter space suggested that infection with the pdmH1N1 virus could reduce either the host’s susceptibility to a subsequent RSV infection, or the infectious period of RSV infection, or RSV infectivity.

The outbreak of pdmH1N1 virus provided an opportunity to investigate viral interference between pdmH1N1 and RSV, as the pdmH1N1 virus emerged as the only circulating influenza virus in the 2009/10 season. With the availability of regional-level data, we were able to dissect variations in the temporal dynamics (i.e., annual/biennial patterns) of RSV activity in different regions in the US and estimate the interaction parameters between the viruses. The increased intensity of the 2010/11 RSV season could be attributed to an increased proportion of susceptible individuals in the population. Similarly, other studies have shown that population susceptibility to influenza and RSV infections increased during the COVID-19 pandemic, leading to larger outbreaks following the relaxation of NPIs [22,23].

We note that none of the mechanisms considered in the model could capture the shift in timing of the 2010/11 RSV epidemic. One possible explanation would be that we only assumed a transient viral interference interval lasting up to a week, occurring in co-infected individuals and disappearing when the infection resolves. Although previous studies have suggested that cross-protection following influenza infection against RSV could last more than two weeks [24], this was not demonstrated in a ferret model, which shares several similarities with the respiratory tracts of humans [25]. The duration of cross-protection between RSV and the pdmH1N1 virus only lasted a week in ferrets [12].

Delayed RSV activity in the 2010/11 season could also be explained by the circulation of other respiratory viruses, which our model did not explicitly include. Our specific emphasis was on studying viral interference from the pdmH1N1 virus on RSV infection, where the interactions are evident at the host level. This focus was guided by experimental studies indicating that the genetic strains of influenza viruses elicit varying levels of host immunity [26]. We also assumed that the RSV-influenza interaction was unidirectional during the 2009/10 season, meaning that only the pdmH1N1 virus would exhibit interference on RSV. This assumption is justified considering that the influenza pandemic preceded the normal 2009/10 RSV season, and our analysis focused on the disruption in RSV activity following the 2009 influenza pandemic. Note that neither could, nor did we intend to show the absence of viral interference from RSV against influenza viruses. To study viral interference from RSV in shaping the transmission dynamics of influenza may require time-series data from multiple epidemic seasons [18]. In addition, we could not differentiate RSV-A and RSV-B in the model due to the data limitation. This may also explain the delayed RSV epidemic in the 2010/11 season.

Dynamic time warping (DTW) is a widely used statistical algorithm [27,28], but its application in identifying various disease transmission patterns has been limited. Recently, multiple studies have used DTW to analyze the trajectories of COVID-19 in different countries, aiming to identify, cluster and predict future trends in disease transmission [29–31]. Here, we utilized DTW and successfully identified the biennial pattern of RSV epidemics in certain regions of the US and atypical RSV seasons following the pandemic. The graphical representation of clusters based on DTW provides an accessible and interpretable method for comparing both temporal and spatial time-series of incidence data, enhancing our understanding of disease transmission patterns longitudinally or geographically. The DTW method also has a promising potential for detecting and identifying atypical epidemic seasons.

We identified three plausible viral interference mechanisms that could shape RSV epidemics following the influenza pandemic. We do acknowledge that these mechanisms are not mutually exclusive. This raises the question of what kind of data would be needed to further distinguish among the models and examine the relative importance of each mechanism for RSV transmission. One possible direction would be to emphasize the incidence of coinfections over a period of time. By fitting the prevalence of coinfections to mathematical models, we could estimate essential kinetic parameters separately, such as the force of infection or the recovery rate from the coinfected components. The comparison between these estimated parameter values and the baseline values (i.e., the rates from individuals infected with one virus) helps discern different mechanisms and evaluate the relative contribution of each process.

There are some limitations to our study. First, we did not have age information on the positive tests for RSV and influenza. Therefore, we assumed a well-mixed population and did not account for varying levels of immunity across different age groups beyond the age-specific contract matrices. It is possible that influenza virus stimulates a weaker innate immune response in young children compared to other age groups [32]. Although our model cannot be used to assess the strength of viral interference in each age group, our results are still sufficient to demonstrate the presence of viral interaction across the entire population. Another limitation is that our model estimates did not show strong viral interference effects between the viruses in Region 4. The difference in the estimates of viral interference for Region 4 is likely attributable to the different pdmH1N1 activity in this region. In regions 1 and 10, there was a strong second wave of pdmH1N1 before the 2009/10 RSV season, whereas the second pdmH1N1 wave was not observed in Region 4. It is not clear why the pandemic influenza exhibited different activity in these regions.

Our findings, which indicate an association between the incidence of RSV infections and pdmH1N1 infections, also have implications for enhanced surveillance of disease transmission of other viruses. As other respiratory viruses (e.g., seasonal influenza viruses and SARS-CoV-2) are expected to co-circulate in the upcoming epidemic seasons, our study provides a framework for studying viral interactions and understanding transmission dynamics. Additional information on the frequency of RSV and other viruses, along with coinfections, would enable us to further validate our results. The mechanistic model proposed in this work is flexible to incorporate the effects of vaccination in preventing disease, such as introducing a model compartment that is resistant to infection and becomes susceptible over time. The extended model can be used for the evaluation of various vaccination scenarios, assessing the impact of vaccination coverage on long-term disease patterns while considering the presence of viral interference. Through systematic analysis of these scenarios, our model can provide valuable insights into the dynamic interplay between vaccination strategies and the patterns of disease transmission, contributing to informed decision-making in public health interventions.

## Data Availability

All data produced in the present work are contained in the manuscript.

## Acknowledgments

This work was supported by a grant from the National Institutes of Health (R01AI137093). The content is solely the responsibility of the authors and does not necessarily represent the official views of the National Institutes of Health.

## Competing interests

DMW has been principal investigator on grants from Pfizer and Merck to Yale University for work unrelated to this manuscript and has received consulting and/or speaking fees from Pfizer, Merck, and GSK/Affinivax. The other authors declare no competing interests.

## Data and availability

All visualization was performed in R (version 4.0.2), and code to produce all figures and data used in this analysis are available in the GitHub repository https://github.com/keli5734/viral-interference.

## Notes

### Competing Interest Statement

The authors have declared no competing interest.

### Author Declarations

The study used (or will use) ONLY openly available human data that were originally located at: https://data.cdc.gov/Laboratory-Surveillance/Respiratory-Syncytial-Virus-Laboratory-Data-NREVSS/52kb-ccu2/about_data

### Summary of Updates

1) Figure 3 and Figure 4 2) abstract

